# Association between Liver Fibrosis and Incident Dementia in the UK Biobank Study

**DOI:** 10.1101/2022.02.09.22270729

**Authors:** Neal S. Parikh, Hooman Kamel, Cenai Zhang, Sonal Kumar, Russell Rosenblatt, Pascal Spincemaille, Ajay Gupta, David E. Cohen, Mony J. de Leon, Rebecca F. Gottesman, Costantino Iadecola

## Abstract

**Background:** There is growing recognition of the role of chronic liver disease in brain health, but the impact of liver fibrosis on dementia risk was unclear. We evaluated the association between liver fibrosis and incident dementia using data from a large prospective cohort study.

**Methods:** We performed a cohort analysis using data from the UK Biobank study, which prospectively enrolled approximately 500,000 adults starting 2007 and continues to follow them. Liver fibrosis was defined using validated cutoffs of the Fibrosis-4 (FIB-4) liver fibrosis score. The primary outcome was incident dementia, ascertained using a validated approach based on participants’ hospital record and mortality data. Secondary outcomes were Alzheimer’s disease and vascular dementia. We excluded participants with prevalent dementia. We used Cox proportional hazards models to evaluate the association between liver fibrosis and incident dementia while adjusting for potential confounders. In additional models, the FIB-4 score was used as a continuous independent variable. Prespecified interaction analyses tested for effect modification by sex, metabolic syndrome, and apolipoprotein E4 carrier status.

**Results:** Among 455,226 participants included in this analysis, the mean age was 56.5 years and 54% were women. Standard liver chemistries were largely in the normal range. However, 2.17% (95% CI, 2.13-2.22%) had liver fibrosis based on their FIB-4 score. The rate of dementia per 1,000 person-years was 1.76 (95% CI, 1.50-2.07) in participants with liver fibrosis and 0.52 (95% CI, 0.50-0.54) in those without. After adjusting for demographics, socioeconomic deprivation, educational attainment, metabolic syndrome, hypertension, diabetes, dyslipidemia, and tobacco and alcohol use, liver fibrosis was associated with an increased risk of dementia (HR, 1.52; 95% CI, 1.22-1.90). An independent association was also noted between FIB-4, treated as a continuous variable, and dementia (HR per unit, 1.29; 95% CI, 1.20-1.39). Effect modification by sex, metabolic syndrome, and apolipoprotein E4 carrier status was not seen.

**Conclusion:** Liver fibrosis in middle age was associated with an increased risk of incident dementia, independent of metabolic syndrome and shared risk factors. Liver fibrosis may be an underrecognized risk factor for dementia.

## Introduction

Common pathologies associated with cognitive impairment, such as those characteristic of Alzheimer’s disease and related dementias, incompletely explain the variance in age-related cognitive decline.^1^ There is growing appreciation of the multiplicity of pathogenic factors underlying cognitive impairment,^2^ and recognition that medical comorbidities may contribute to cognitive impairment and dementia.^3^

The liver-brain axis is a burgeoning area of investigation, underpinned by mechanistic and epidemiological studies that substantially expand the scope of liver-brain pathways beyond ammonia clearance.^4-6^ Chronic liver disease occurs along a spectrum, with liver fibrosis occurring in response to chronic liver injury from a variety of causes.^7^ Apart from people with clinically apparent cirrhosis, which is the end stage of liver fibrosis, individuals with liver fibrosis commonly have normal liver enzymes and lack clinical signs and symptoms typically associated with liver disease.^8-12^ Even in this subclinical stage, liver fibrosis is associated with cardiovascular disease.^13-17^

There are few data on the relationship between liver fibrosis and brain health at the population level. We previously demonstrated the cross-sectional association between liver fibrosis and worse cognitive performance in a population-based sample.^18^ However, longitudinal data regarding incident dementia are lacking. Therefore, using data from a large, prospective cohort, we evaluated the hypothesis that liver fibrosis is associated with an increased risk of future dementia.

## Methods

### Design

This is a retrospective cohort study using data from the United Kingdom (UK) Biobank. The UK Biobank is a prospective cohort study that recruited approximately 500,000 participants between 40-69 years of age from across the UK.^19^ In the UK, nearly all people are registered with a general practitioner through the National Health Service; eligible participants were identified from National Health Service registers based on age and residence within reasonable traveling distance of one of 35 assessment centers around the UK. Participants were sent invitation letters by post. Baseline assessments occurred from 2007 to 2010, and included detailed health questionnaires, physical measurements, and collection of biological samples for laboratory panels and genotyping. Standard laboratory panels included blood count and blood biochemistry tests, such as those required for this analysis. Participants provided signed, informed consent to be followed longitudinally, including through linked hospital record and death data. Based on these linked data, the UK Biobank includes time-to-event data for common health outcomes, including dementia, in addition to death and loss to follow-up. UK Biobank data, including externally collected data such as hospital record and death registry data, are subjected to extensive data stewardship measures designed to ensure data integrity.^20^ The data that support the findings of this study are made available in an anonymized format to qualified investigators upon application to the UK Biobank. Analytic methods will be made available upon reasonable request. The Weill Cornell Medicine institutional review board certified these analyses of deidentified data as exempt from review.

### Population

From among the approximately 500,000 people in the UK Biobank, we included all participants who attended a baseline assessment center visit while excluding participants with prevalent dementia, missing exposure variable data, possible acute hepatitis, and severe thrombocytopenia. Participants who had prevalent dementia were identified based on a dementia diagnosis concurring with or preceding baseline assessment. We excluded people with possible acute hepatitis (aspartate aminotransferase or alanine aminotransferase ≥250 international units/liter),^21^ and people with severe thrombocytopenia (<50,000 per microliter)^22^ to minimize the influence of these conditions on our findings through impact on the liver fibrosis score. These exclusions resulted in a study sample of 455,226 participants (eFigure1 in the Supplement).

### Measurements

The exposure variable was liver fibrosis. We calculated the Fibrosis-4 (FIB-4) score for each participant according to its formula:

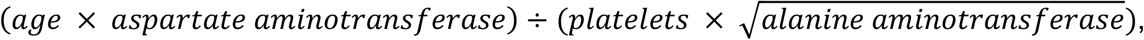

with age in years, aminotransferases in units/liter, and platelet count in 10^9^/liter.^23^ In the primary analysis, we classified participants with FIB-4 score > 2.67 as having liver fibrosis and those with FIB-4 ≤ 2.67 as not having liver fibrosis.^23,24^ A secondary approach treated the FIB-4 score as a continuous variable, including visualization as a restricted cubic spline. The FIB-4 score has been validated to have good accuracy for liver fibrosis across multiple conditions, including nonalcoholic fatty liver disease, alcoholic liver disease, hepatitis B, and hepatitis C.^24-30^ As evidence of its validity in people unselected for individual liver conditions, a high FIB-4 score among the general population was associated with a 17-fold higher risk of future clinically apparent liver disease.^31^ Our approach of evaluating liver fibrosis in the general population, irrespective of underlying etiology, is consistent with other efforts to understand the role of liver fibrosis broadly in human disease.^13,14,32,33^ Furthermore, this approach is consistent with emerging liver fibrosis screening paradigms^8,34^ and accounts for the real-world overlap between chronic liver conditions such as alcoholic and presumed non-alcoholic fatty liver disease.^35^

The primary outcome was incident all-cause dementia. UK Biobank developed algorithms to identify dementia based on medical history, hospital records, and mortality register data using *International Classification of Diseases* versions 9 and 10 diagnosis codes.^36^ The UK Biobank dataset contains time-to-event data for dementia derived using these algorithms for the entire study cohort. In a validation study of UK Biobank data, the positive predictive value of this approach was 85% for all-cause dementia, 71% for Alzheimer’s disease, and 33% for vascular dementia.^37^ Alzheimer’s disease and vascular dementia were included as secondary outcomes in the present analysis.

We tabulated a variety of population characteristics. Demographics included age, sex, self-reported race/ethnicity, socioeconomic status, and educational attainment. Race/ethnicity were categorized as Asian, Black, other/multiple, and White, with categorization reflecting UK demographics. Socioeconomic status at baseline was measured using the Townsend deprivation index,^38^ which incorporates employment status, automobile ownership, home ownership, and household crowding; higher scores indicate worse socioeconomic deprivation. Educational attainment was categorized as professional/university degree, pre-university/secondary education qualifications, vocational training, and certificate of secondary education. Cardiometabolic risk factors were assessed at baseline. Hypertension was defined as self-reported diagnosis of hypertension, systolic blood pressure ≥ 130 mmHg, diastolic blood pressure ≥ 80 mmHg, or use of anti-hypertensive medications. Diabetes was defined as a self-reported diagnosis of diabetes, hemoglobin A1c ≥ 6.5%, or use of diabetes medications. Metabolic syndrome was defined as having at least 3 of the following 5 features: waist circumference ≥ 102 centimeters in men or ≥ 88 centimeters in women, triglycerides ≥ 150 milligrams per deciliter, HDL cholesterol < 40 milligrams per deciliter in men or < 50 milligrams per deciliter for women, systolic blood pressure ≥ 130 mmHg or diastolic blood pressure ≥ 85 mmHg, and serum glucose ≥ 100 milligrams per deciliter. Dyslipidemia was defined as use of lipid-lowering medications or total cholesterol ≥ 200 milligrams per deciliter. BMI was categorized using standard cutoffs for normal (18.5-24.99 kg/m^2^), underweight (<18.5 kg/m^2^), overweight (25-29.99 kg/m^2^), and obesity (≥30 kg/m^2^). Tobacco use was categorized as never smoker versus smoker. Alcohol use was categorized based on consumption frequency as non-drinker, less than once weekly, 1-4 times per week, and daily or almost daily. Patients reporting any prevalent chronic liver condition, including cirrhosis, alcoholic liver disease, hepatitis and non-infectious liver disease during their nurse-led interview were categorized as having a clinically known liver condition. Participants with at least one apolipoprotein E4 allele were categorized as carriers. The apolipoprotein gene was directly genotyped for UK Biobank participants, and we used standard definitions to assign genotypes based on *rs429358* and *rs7412* alleles.^39,40^

### Statistical Analyses

Survival analysis was used to estimate cumulative risks while censoring at the time of death, loss to follow-up, or at the end of available follow-up in our dataset (March 1, 2018). The primary statistical analysis entailed use of Cox proportional hazards models to compare the risk of dementia in people with and without liver fibrosis by calculating cause-specific hazard ratios. Proportionality of hazards was confirmed by visual inspection of survival and log-log plots. In a secondary approach, the FIB-4 score was instead treated as a linear variable to assess the association between each 1-unit increase in FIB-4 and the risk of dementia. FIB-4 was also fit as a restricted cubic spline in order to visualize the relationship with incident dementia. We constructed the following prespecified models. Model 1 was unadjusted. Model 2 was adjusted for age, sex, race/ethnicity, educational attainment, and socioeconomic deprivation. Model 3 was additionally adjusted for hypertension, diabetes, dyslipidemia, body mass index, metabolic syndrome, smoking status, and alcohol consumption frequency. Of note, hypertension and diabetes may be downstream sequelae of nonalcoholic fatty liver disease (a common cause of liver fibrosis)^41,42^ and therefore could be considered as potential mediators in the relationship between liver fibrosis and dementia; however, we included them as potential confounders to derive a conservative estimate of the independent contribution of liver fibrosis to dementia risk.

We performed several sensitivity analyses. First, we replaced hypertension, diabetes, and dyslipidemia with mean systolic blood pressure, hemoglobin A1c, and total cholesterol, respectively. Second, we excluded all 2,652 participants with any known chronic liver condition at baseline to evaluate the association between subclinical liver fibrosis and incident dementia. Third, we restricted the study sample to the 36,835 participants who reported zero alcohol consumption to minimize the risk of confounding by alcohol use. We prespecified several tests of interaction with liver fibrosis in models for the primary outcome of all-cause dementia. First, effect modification by sex was assessed because stronger associations between chronic liver disease and cerebrovascular disease have been reported in women.^43,44^ Second, we evaluated for effect modification by metabolic syndrome. Third, we evaluated whether the association between liver fibrosis and dementia varies based on genetic risk for dementia conferred by apolipoprotein E4 carrier status. Effect modification was evaluated based on the P-value of cross-product terms on the multiplicative scale. The threshold of statistical significance was set at α = 0.05. All statistical analyses were performed using SAS Version 9.4 (Cary, NC), and data were visualized using R 4.0.5 (R Core Team, Vienna, Austria).

## Results

From among 502,613 participants in the UK Biobank, we included 455,226 participants after exclusions (eFigure1 in the Supplement). Missing FIB-4 score data was the most frequent reason for exclusion; participants with and without missing FIB-4 score data appeared similar (eTable1 in the Supplement). The mean age of participants was 56.5 (SD, 8.1) years, and 246,466 (54%) were women. Clinically known liver conditions were uncommon, reported by 2,652 (0.6%) participants. The median FIB-4 score was 1.24 (IQR, 0.97-1.57), falling in the indeterminate range. However, 9,895 (2.2%; 95% CI, 2.1-2.2%) participants had a FIB-4 score consistent with liver fibrosis. Participants with liver fibrosis were older, more likely men, had a higher prevalence of hypertension and diabetes, and more often reported daily alcohol consumption (Table 1).

**Table 1.** Baseline characteristics^a^ of participants in the UK Biobank, stratified by liver fibrosis.

|  | With Liver Fibrosis | Without Liver Fibrosis |
| --- | --- | --- |
| Participants | N=9,895 | N=445,331 |
| Age, years (mean, SD) | 62.5 (5.9) | 56.4 (8.1) |
| Female | 3,334 (34%) | 243,132 (55%) |
| Race/ethnicity<sup>b</sup> |  |  |
| Asian | 179 (2%) | 9,933 (2%) |
| Black | 277 (3%) | 6,637 (2%) |
| Other and multiracial | 122 (1%) | 6,583 (1%) |
| White | 9,243 (94%) | 420,130 (95%) |
| Education<sup>c</sup> |  |  |
| University graduate/Professional | 3,467 (47%) | 167,506 (46%) |
| Pre-university qualifications | 2,791 (38%) | 29,119 (8%) |
| Vocational school completion | 788 (11%) | 144,109 (39%) |
| Certificate of Secondary Education only | 279 (4%) | 24,248 (7%) |
| Socioeconomic deprivation<sup>d</sup> |  |  |
| Quartile 1 (lowest deprivation) | 2,386 (24%) | 112,597 (25%) |
| Quartile 2 | 2,369 (24%) | 111,691 (25%) |
| Quartile 3 | 2,495 (25%) | 111,255 (25%) |
| Quartile 4 (highest deprivation) | 2,644 (27%) | 109,788 (25%) |
| Hypertension | 7,783 (79%) | 320,159 (72%) |
| Systolic blood pressure, mmHg (mean, SD) | 142 (20) | 138 (19) |
| Diabetes | 1,035 (10%) | 25,973 (6%) |
| Hemoglobin A1c, % (mean, SD) | 5.5 (0.7) | 5.5 (0.6) |
| Dyslipidemia | 7,124 (72%) | 332,933 (75%) |
| Total cholesterol, mg/dL (mean, SD) | 205 (46) | 220 (43) |
| History of tobacco smoking | 6,239 (63%) | 265,014 (60%) |
| Metabolic syndrome | 2,918 (29%) | 125,639 (28%) |
| Body mass index, kg/m<sup>2</sup> (mean, SD) | 27.3 (4.9) | 27.4 (4.8) |
| Known liver condition | 228 (2.3%) | 2,424 (0.5%) |
| Alcohol consumption frequency |  |  |
| None | 969 (9.8%) | 35,866 (8.1%) |
| Less than once weekly | 1,910 (19.3%) | 100,764 (22.7%) |
| 1-4 times weekly | 4,241 (43.0%) | 218,303 (43.0%) |
| Daily or almost daily | 2,755 (27.9%) | 89,893 (20.2%) |
| Aspartate aminotransferase, IU/L (median, IQR) | 34 (27-49) | 24 (21-29) |
| Alanine aminotransferase, IU/L (median, IQR) | 22 (15-34) | 20 (15-27) |
| Platelet count, 10<sup>9</sup> cells/liter (median, IQR) | 155 (127-182) | 250 (216-288) |
| Albumin, g/dl (median, IQR) | 4.5 (4.3-4.7) | 4.5 (4.4-4.7) |

Abbreviations: SD, standard deviation; mmHg, millimeters mercury; mg/dL, milligrams per deciliter; kg/m<sup>2</sup>, kilograms per meter-squared; IU/L, international units per liter; IQR, interquartile range; g/dl, grams per deciliter.
<sup>a</sup>Data are presented as percentage (95% confidence interval of percentage) unless otherwise specified.
Percentages for any given characteristic may not sum to 100% because of rounding.
<sup>b</sup>Race/ethnicity groupings based on UK Biobank population composition.
<sup>c</sup>Education categorized based on UK educational system. A and O level certifications indicate higher educational attainment than Certificate of Secondary Education.
<sup>d</sup>Socioeconomic deprivation measured using the Townsend deprivation index, reflecting employment status, automobile ownership, home ownership, and household crowding; higher scores indicate worse socioeconomic deprivation.

During a median follow-up of 9.0 (IQR, 8.3-9.7) years, 2,223 cases of incident all-cause dementia occurred. The rate of all-cause dementia per 1,000 person-years was 1.76 (95% CI, 1.50-2.07) in participants with liver fibrosis and 0.52 (95% CI, 0.50-0.54) in those without (Figure 1). In an unadjusted Cox proportional hazards model, participants with liver fibrosis had an increased risk of incident dementia as compared to participants without liver fibrosis (hazard ratio [HR], 3.45; 95% CI, 2.92-4.08). This association was attenuated and remained significant in adjusted models, including when adjusting for demographics, education, and shared risk factors for liver fibrosis and dementia (HR, 1.52; 95% CI, 1.22-1.90) (Table 2). Findings were similar when modeling the FIB-4 score as a continuous variable (adjusted HR per unit, 1.29; 95% CI, 1.20-1.39) (Table 2), and visualization of the FIB-4 score as a restricted cubic spline corroborated this relationship (Figure 2).

**Table 2.** Association^a^ of Liver Fibrosis with Incident Dementia in the UK Biobank.

| **Model** | **Liver Fibrosis versus**<br>**no Liver Fibrosis** | **For each 1.0 unit**<br>**increase in FIB-4** |
| --- | --- | --- |
| Model 1: Unadjusted | 3.45 (2.92-4.08) | 1.60 (1.56-1.64) |
| Model 2: Adjusted for sociodemographics<sup>b</sup> | 1.60 (1.29-1.99) | 1.32 (1.23-1.42) |
| Model 3: Adjusted for sociodemographics, comorbidities<sup>c</sup> | 1.52 (1.22-1.90) | 1.29 (1.20-1.39) |
| Sensitivity analysis 1<sup>d</sup> | 1.51 (1.19-1.91) | 1.30 (1.20-1.40) |
| Sensitivity analysis 2<sup>e</sup> | 1.45 (1.16-1.83) | 1.26 (1.17-1.36) |
| Sensitivity analysis 3<sup>f</sup> | 1.96 (1.12-3.43) | 1.27 (1.04-1.55) |

Abbreviations: FIB-4, Fibrosis-4 score.
<sup>a</sup>Reported as hazard ratio (95% confidence interval).
<sup>b</sup>Age, sex, race/ethnicity, educational attainment, and socioeconomic deprivation.
<sup>c</sup>Hypertension, diabetes, dyslipidemia, body mass index, metabolic syndrome, smoking, and alcohol intake frequency.
<sup>d</sup>Systolic blood pressure, hemoglobin A1c, and total cholesterol were used as continuous covariates in place of hypertension, diabetes, and dyslipidemia, respectively.
<sup>e</sup>Excluded participants with any clinically known liver condition at baseline.
<sup>f</sup>Excluded participants with any non-zero alcohol consumptions at baseline.

**Figure 1.**
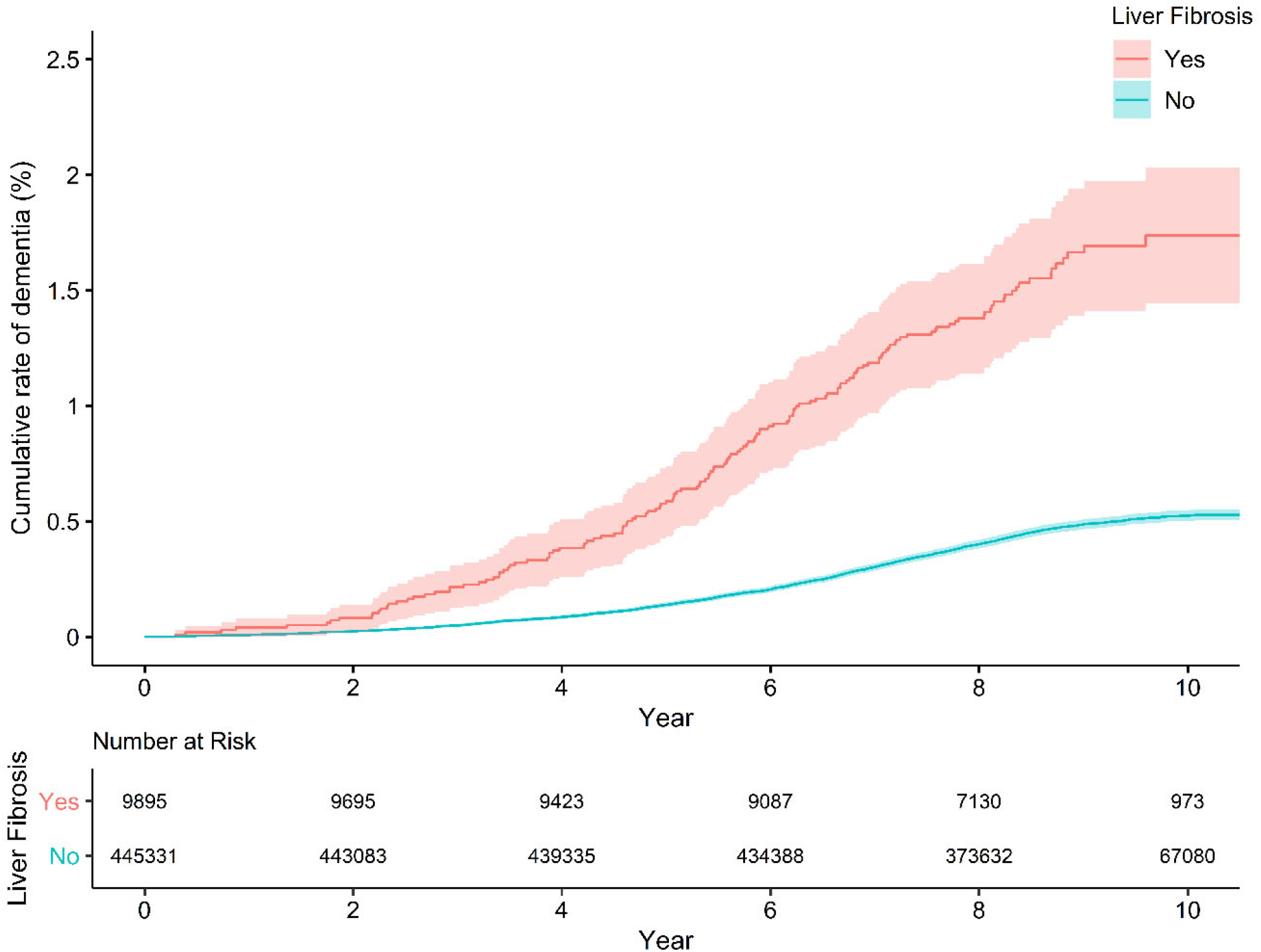
Cumulative risk of incident all-cause dementia, stratified by liver fibrosis. In this analysis of data from the UK Biobank prospective cohort study, participants with liver fibrosis had an increased risk of all-cause dementia.

**Figure 2.**
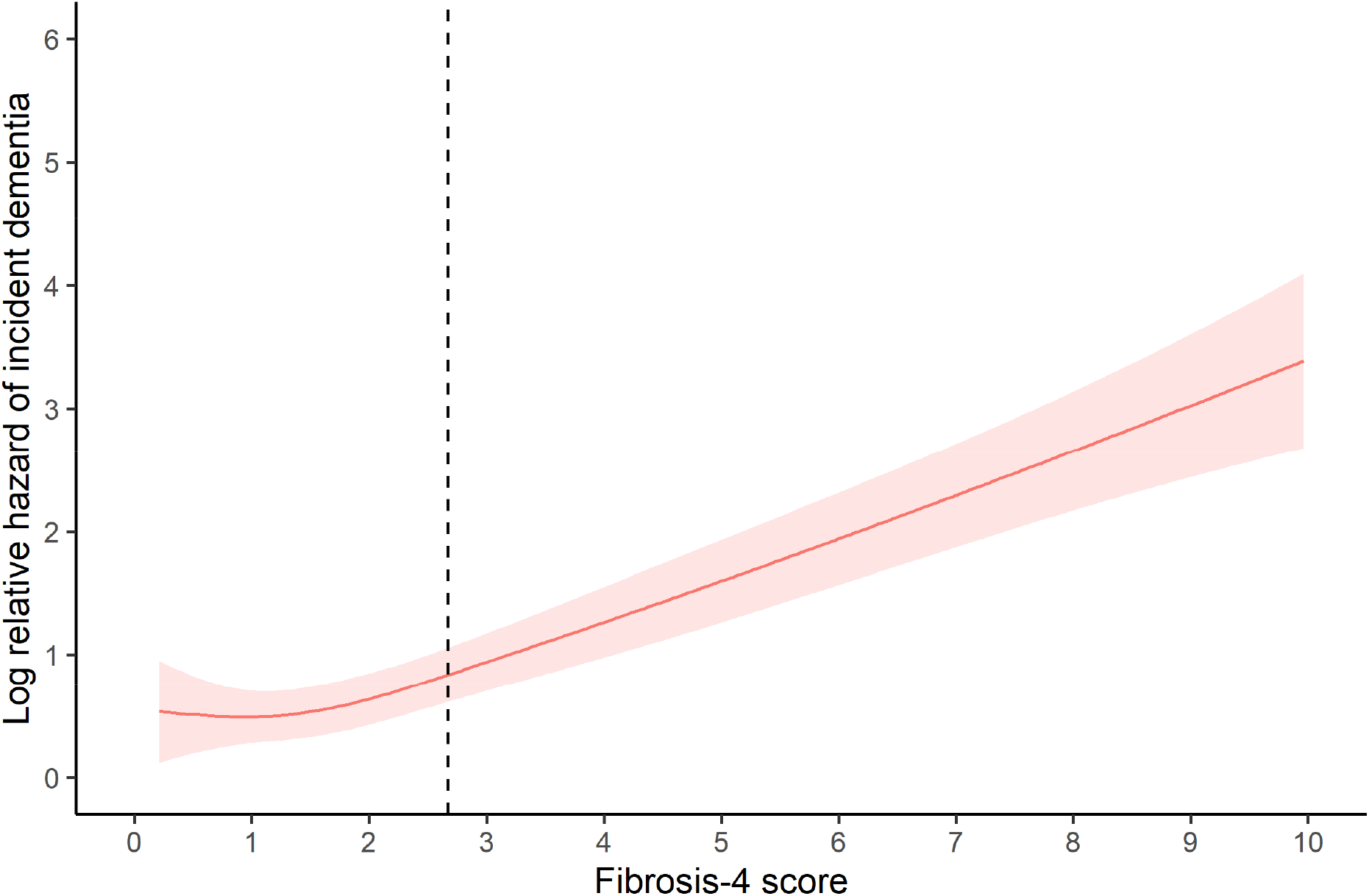
Relative Hazard of Incident Dementia by Fibrosis-4 Score as a Restricted Cubic Spline. This restricted cubic spline visualization displays hazards of dementia relative to a reference Fibrosis-4 score of 2.67, which is the cut-point used to define liver fibrosis in the primary analysis. The model was adjusted for age, sex, race/ethnicity, educational attainment, socioeconomic deprivation, hypertension, diabetes, dyslipidemia, body mass index, metabolic syndrome, smoking, and alcohol intake frequency. The shaded areas represent the 95% confidence interval band.

Our findings were unchanged in a sensitivity analysis that adjusted for systolic blood pressure, hemoglobin A1c, and total cholesterol in place of corresponding categorical variables (Table 2). In additional sensitivity analyses, results were consistent after excluding participants with any clinically known liver conditions at baseline and, separately, when restricting the study sample to participants reporting zero alcohol consumption at baseline (Table 2). In formal tests of interaction, the association between FIB-4 score, as a continuous variable, and incident dementia was not modified by sex (P=0.90), metabolic syndrome (P=0.23), or apolipoprotein E4 carrier status (P=0.25). There was also no effect modification by sex (P=0.63), metabolic syndrome (P=0.10), or apolipoprotein E4 carrier status (P=0.88) in models that categorized participants as having liver fibrosis based on FIB-4 score.

In terms of secondary outcomes, there were 836 cases of incident Alzheimer’s disease and 467 cases of vascular dementia. Liver fibrosis was non-significantly associated with a higher risk of Alzheimer’s disease in adjusted models when categorizing participants based on the FIB-4 score (HR, 1.31; 95% CI, 0.89-1.93), and significantly associated when treating the FIB-4 score as a continuous variable (HR per unit, 1.24; 95% CI, 1.09-1.41). Liver fibrosis was significantly associated with an increased risk of vascular dementia in adjusted models both when treating the FIB-4 score as a categorical variable (HR, 1.64; 95% CI, 1.03-2.60) and continuous variable (HR, 1.24; 95% CI, 1.05-1.46).

## Discussion

In this analysis of adults in the UK Biobank cohort study, participants with liver fibrosis based on the FIB-4 liver fibrosis score had an increased risk of incident dementia independently of shared risk factors. This finding was robust to multiple sensitivity analyses, for example when excluding participants with any clinically known liver conditions or alcohol use.

Our findings provide new evidence that liver fibrosis is associated with a future risk of dementia at the population level. Prior studies of liver fibrosis and cognition have involved cross-sectional study designs or highly selected cohorts. For example, we previously found a cross-sectional association between liver fibrosis and cognitive performance in a population-based cohort of older Americans in the National Health and Nutrition Examination Survey, which lacks longitudinal dementia outcomes data.^18^ Other cross-sectional studies identified an association of liver fibrosis with worse cognitive performance in narrowly defined populations, such as in people with human immunodeficiency virus and hepatitis C virus infection or people with computed tomography evidence of hepatic steatosis.^45-48^ In this context, our study adds novel evidence that liver fibrosis is associated with worse future cognitive health in the population at large. Furthermore, this association was observed in a population with a mean age of 56.5 years at cohort entry, adding liver fibrosis to a growing list of midlife risk factors for dementia.^49^

There are several possible explanations for our findings. Hepatic encephalopathy is common in individuals with end-stage cirrhosis.^50-52^ Some individuals with liver fibrosis may have had subclinical cirrhosis or later progressed to develop clinically apparent cirrhosis. Thus, some cases of incident dementia may have partly reflected an underlying hepatic encephalopathy. For example, hepatic encephalopathy may have unmasked underlying cognitive impairment, and some participants with hepatic encephalopathy may have been misdiagnosed as having dementia. However, hepatic encephalopathy is typically a well-recognized syndrome unlikely to be classified as dementia, and our findings were unchanged when we excluded people with known liver conditions in a sensitivity analysis. Apart from hepatic encephalopathy, there are several additional mechanistic links between liver fibrosis and dementia. Liver fibrosis may promote the development and progression of vascular brain injury, as suggested in prior cross-sectional studies.^53,54^ We found an association between liver fibrosis and dementia after adjusting for vascular risk factors; shared vascular risk factors alone do not explain our findings. There is also growing evidence supporting the role of the liver in peripheral amyloid-β clearance, with abnormal liver function potentially worsening cerebral amyloid-β deposition.^55-58^ Lastly, the liver may be responsible for generating neuroprotective hepatokines,^59^ although the implications of liver fibrosis on such processes is not clear. Future studies should take advantage of emerging brain imaging and plasma-based biomarkers of brain health to evaluate the relative contributions of these processes.

The key strengths of this analysis are the large sample size, use of a validated liver fibrosis score, and follow-up of sufficient time to assess the impact of liver fibrosis in midlife on an age-related incident outcome. The findings should be interpreted considering several limitations. First, while validated, the FIB-4 liver fibrosis score is not a direct measure of liver fibrosis, and the use of a surrogate marker introduces the possibility of misclassification. Participants with mild liver fibrosis may be misclassified as having no fibrosis, for example. The consistency of our findings, whether categorizing participants based on FIB-4 scores or using FIB-4 score as continuous variable, partially mitigates the impact of misclassification introduced by cut-offs. Regardless, future studies should use more direct liver measures such as transient elastography or magnetic resonance elastography, although the feasibility of this is limited in cohorts of this size. Second, the UK Biobank does not include Alzheimer’s disease biomarkers, and lacks liver-related data such as hepatokine levels and serum ammonia levels. Additionally, dementia diagnoses are ascertained based on a validated UK Biobank outcome algorithm, and this lacks specificity for Alzheimer’s disease, vascular dementia, and other specific forms of dementia. Future studies should incorporate more neurocognitive data or adjudicated dementia outcome data to determine which dementia type is most impacted by the liver. Such efforts may also uncover mechanisms that underlie the link between liver fibrosis and dementia. Third, the UK Biobank study population is not representative of the UK population given the evidence of a healthy participant bias.^60^ In this analysis, the prevalence of liver fibrosis was approximately 2.2%, which is consistent with but on the lower range of estimates from other population-based studies.^61^ For example, in our prior analysis of older Americans in a population-based cohort, approximately 5% had liver fibrosis by the same criteria employed in this analysis.^18^ The UK Biobank population is also not representative of other countries with different racial and ethnic compositions, health-related behavior patterns, and prevalent medical conditions. Therefore, our results may not be generalizable to the population at large. Future studies should answer whether liver fibrosis impacts brain health among more diverse populations, and among populations with a greater burden of comorbidities.

## Conclusion

Liver fibrosis in midlife was associated with an increased future risk of dementia, including after accounting for shared risk factors and when excluding participants with clinically known liver conditions at baseline. Liver fibrosis may be an important determinant of long-term brain health outcomes.

## Data Availability

This research has been conducted using the UK Biobank Resource. The data that support the findings of this study are made available in an anonymized format to qualified investigators upon application to the UK Biobank. Analytic methods will be made available upon reasonable request.

https://www.ukbiobank.ac.uk/

## Authors’ contributions

Dr. Parikh was responsible for conception and design of the work, analysis and interpretation of the data, drafting of the work, and giving final approval of the version to be published; and, he agrees to be accountable for all aspects of the work in ensuring that questions related to the accuracy or integrity of any part of the work are appropriately investigated and resolved. Ms. Zhang was responsible for analysis and interpretation of the data, revising the work critically for important intellectual contents, and providing final approval of the version to be published; and, she agrees to be accountable for all aspects of the work in ensuring that questions related to the accuracy or integrity of any part of the work are appropriately investigated and resolved. Drs. Kumar, Rosenblatt, Spincemaille, and Gupta were responsible for interpretation of the data, revising the work critically for important intellectual content, and providing final approval of the version to be published; and, they agree to be accountable for all aspects of the work in ensuring that questions related to the accuracy or integrity of any part of the work are appropriately investigated and resolved. Drs. Kamel, de Leon, Gottesman, and Iadecola were responsible for design of the work, interpretation of the data, revising the work critically for important intellectual content, providing supervision, and giving final approval of the version to be published; and, they agree to be accountable for all aspects of the work in ensuring that questions related to the accuracy or integrity of any part of the work are appropriately investigated and resolved.

## Data Access, Reproducibility, and Analysis

Dr. Parikh and Ms. Zhang had full access to all the data in the study and take responsibility for the integrity of the data and the accuracy of the data analysis.

