## Supplemental Materials for "Association between Liver Fibrosis and Incident Dementia in the UK Biobank Study"

**Contents**:

Supplemental Table I. Baseline characteristics of participants in the UK Biobank, stratified by missingness of Fibrosis-4 liver fibrosis score.

Supplemental Figure 1. Study population flow chart.

**Supplemental Table I.** Baseline characteristics[a] of participants in the UK Biobank, stratified by missingness of Fibrosis-4 liver fibrosis score.

| | **Missing** | **Not missing** |
|---|---|---|
| Participants | N=46,774 | N=455,839 |
| Age, years (mean, SD) | 56.4 (8.2) | 56.5 (8.1) |
| Female | 26,599 (57%) | 246,754 (54%) |
| Race/ethnicity[b] | | |
| Asian | 1,317 (3%) | 10,136 (2%) |
| Black | 1,137 (2%) | 6,923 (2%) |
| Other and multiracial | 805 (2%) | 6,709 (1%) |
| White | 42,710 (93%) | 429,946 (95%) |
| Education[c] | | |
| University graduate/Professional | 15,786 (46%) | 17,1160 (46%) |
| Pre-university qualifications | 13,410 (39%) | 147,097 (39%) |
| Vocational school completion | 2,766 (8%) | 29,958 (8%) |
| Certificate of Secondary Education only | 2,328 (7%) | 24,559 (7%) |
| Socioeconomic deprivation[d] | | |
| Quartile 1 (lowest deprivation) | 11,067 (24%) | 115,118 (25%) |
| Quartile 2 | 11,338 (24%) | 114,194 (25%) |
| Quartile 3 | 11,536 (25%) | 113,908 (25%) |
| Quartile 4 (highest deprivation) | 12,833 (27%) | 112,619 (25%) |
| Hypertension | 32,495 (69%) | 328,420 (72%) |
| Diabetes | 2,971 (6%) | 27,066 (6%) |
| Dyslipidemia | 13,280 (28%) | 340, 482 (75%) |
| History of tobacco smoking | 27,137 (58%) | 271,612 (60%) |
| Metabolic syndrome | 12,134 (26%) | 128,758 (28%) |
| Body mass index, $kg/m^2$ (mean, SD) | 27.7 (5.1) | 27.4 (4.8) |
| Known liver condition | 244 (2%) | 2,426 (1%) |
| Alcohol consumption frequency | | |
| None | 4,767 (10.2%) | 36,919 (8.1%) |
| Less than once weekly | 11,035 (23.6%) | 102,814 (22.6%) |
| 1-4 times weekly | 21,922 (46.9%) | 222,788 (48.9%) |
| Daily or almost daily | 8,974 (19.2%) | 92,791 (20.4%) |
| Aspartate aminotransferase, IU/L (median, IQR) | 24 (21-29) | 24 (21-29) |
| Alanine aminotransferase, IU/L (median, IQR) | 19 (15-27) | 20 (15-27) |
| Platelet count, $10^9$ cells/liter (median, IQR) | 245 (210-284) | 248 (213-287) |
| Albumin, g/dl (median, IQR) | 4.5 (4.3-4.7) | 4.5 (4.3-4.7) |

Abbreviations: SD, standard deviation; $kg/m^2$, kilograms per meter-squared; IU/L, international units per liter; IQR, interquartile range; g/dl, grams per deciliter.

[a]Data are presented as percentage (95% confidence interval of percentage) unless otherwise specified. Percentages for any given characteristic may not sum to 100% because of rounding.

[b]Race/ethnicity groupings based on UK Biobank population composition.

[c]Education categorized based on UK educational system. A and O level certifications indicate higher educational attainment than Certificate of Secondary Education.

[d]Socioeconomic deprivation measured using the Townsend deprivation index, reflecting employment status, automobile ownership, home ownership, and household crowding; higher scores indicate worse socioeconomic deprivation.

**Supplemental Figure I.** Study population flow chart.

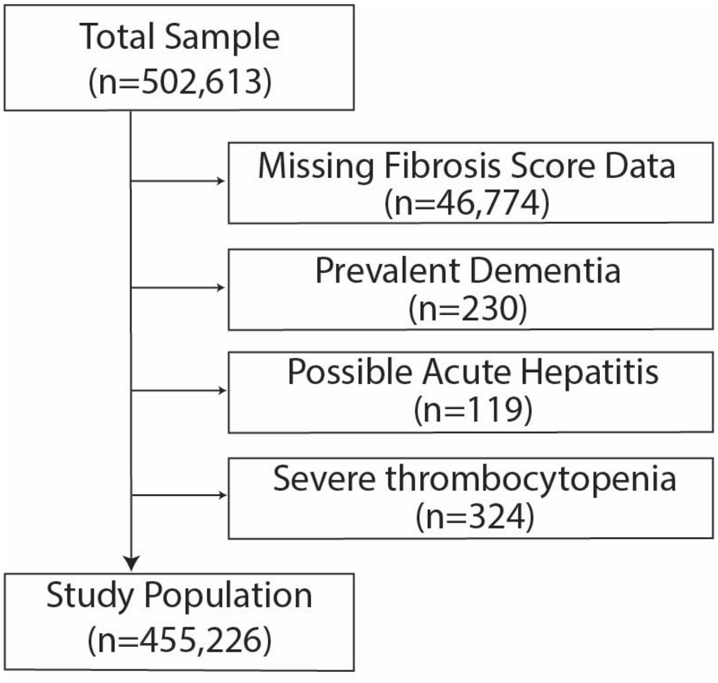


**Legend**: We excluded participants with missing liver fibrosis score data, prevalent dementia, possible acute hepatitis (aspartate or alanine aminotransferase $\geq$ 250 international units per liter), and severe thrombocytopenia (platelet count <50,000 per microliter). Some participants had multiple reasons for exclusion.
